# Multicenter normative data for mesopic microperimetry

**DOI:** 10.1101/2024.02.05.24302327

**Authors:** Maximilian Pfau, Jasleen K. Jolly, Jason Charng, Leon von der Emde, Philipp L. Müller, Georg Ansari, Kristina Pfau, Fred K Chen, Zhichao Wu

## Abstract

**Purpose:** To provide a large, multi-center normative dataset for the Macular Integrity Assessment (MAIA) microperimeter and compare the goodness-of-fit and prediction interval calibration-error for a panel of hill-of-vision models.

**Methods:** Microperimetry examinations from five independent study groups and one previously available dataset were included. Linear mixed models (LMMs) were fitted to the data to obtain interpretable hill-of-vision models. For predicting age-adjusted normative values, an array of regression models were compared using cross-validation with site-wise splits. The mean absolute error (MAE) and miscalibration area (area between the calibration curve and the ideal diagonal) were evaluated as the performance measures.

**Results:** 1,052 tests from 531 eyes of 432 participants were included. Based on the parameters ‘participant age’, ‘eccentricity from the fovea’, ‘overlap with the central fixation target’ and ‘eccentricity along the four principal meridians’, a Bayesian mixed model had the lowest MAE (2.13 dB; 95% confidence interval [CI] = 1.86, 2.40 dB) and miscalibration area (0.14; 95% CI = 0.07, 0.20). However, a parsimonious linear model provided a comparable MAE (2.16 dB; 95% CI = 1.89, 2.43 dB) and a similar miscalibration area (0.14; 95% CI = 0.08, 0.20).

**Conclusions:** Normal variations in visual sensitivity on mesopic microperimetry can be effectively explained by a linear model that includes age and eccentricity. The dataset and a code vignette are provided for estimating normative values across a large range of retinal locations, applicable to customized testing patterns.

## INTRODUCTION

Best-corrected visual acuity (BCVA) is the most commonly used method to assess visual function in clinical trials.^1^ It is suitable for tracking the progression of diseases that imminently threaten foveal vision. However, in retinal conditions where there is either extensive foveao-macular atrophy or foveal-sparing atrophy, BCVA may remain stable at a low or high level respectively for a prolonged duration.^2^ In such conditions, including geographic atrophy (GA) secondary to age-related macular degeneration (AMD) or inherited retinal diseases, central visual field measurement maybe more suitable for assessing disease progression.^3^

Microperimetry, also known as fundus-controlled perimetry, is a technique that allows for retinotopic mapping of light sensitivity, even in patients with unstable fixation.^3^ This technique has been used as a clinical trial outcome measure, particularly in the form of mesopic white-on-white microperimetry.^4,5^ Whilst microperimetry testing is often performed with standardized stimulus patterns, customized and spatially-specific testing tailored to the disease – such as those sampling regions at high risk of disease progression – could allow functional changes to be more effectively characterized and monitored over time. This has been performed for instance using patient-tailored stimulus patterns that predominantly sample the perilesional regions in eyes with GA,^6^ or high-density targeted testing of early atrophic lesions in AMD.^7–9^ Furthermore, analyses of the results from microperimetry testing with standardized stimulus patterns could also be limited *post hoc* to at-risk locations, such as those in the junctional zone external to atrophic regions in eyes with GA^10,11^ and Stargardt disease,^12,13^ or those internal to the boundary of atrophic region in retinitis pigmentosa.^14^

To accurately interpret microperimetry (or any type of perimetry) data, it is critical to determine the sensitivity loss from the expected age- and location-specific normal sensitivity.^15^ In addition, there are specific regions of the visual field where the interindividual variation in visual sensitivity may be lower or higher.^16^ Thus, the determination of pathological visual sensitivity loss relies on the knowledge of the age- and location-dependent normative sensitivity and interindividual variability; both of which are challenging to measure especially with customized or patient-tailored microperimetry testing due to the individualized nature of the test locations.^6,7^

To date, most work seeking to establish such normative data for microperimetry is based on separate analyses of a pre-defined test locations from standardized stimulus patterns.^17,18^ One previous publication by Denniss and Astle^19^ proposed using spatial interpolation to obtain normative data from specific retinal locations. Importantly, the authors provided a first public dataset.^20^ However, their data covered only a limited age range (with all but one participant being between 19 to 31 years old) and it was only obtained from one site and device. A systematic comparison of statistical modeling approaches for normative sensitivity and variability was also not explored in their study.

This study thus aims to (1) provide a comprehensive, multicenter, normative dataset for mesopic microperimetry with a widely used device, (2) systematically compare regression models regarding goodness-of-fit, marginal coverage and conditional coverage, and (3) assess the benefit of limited site-specific normative data.

## METHODS

This study includes data from healthy individuals examined across five centers (participants described in Table 1, microperimetry grids described in Supplementary Table S1 and Figure 1). The data collection adhered to the Declaration of Helsinki, and the study protocols were reviewed by the respective ethics committees. All participants were informed of the nature of the study and provided written informed consent.

**Figure 1.**
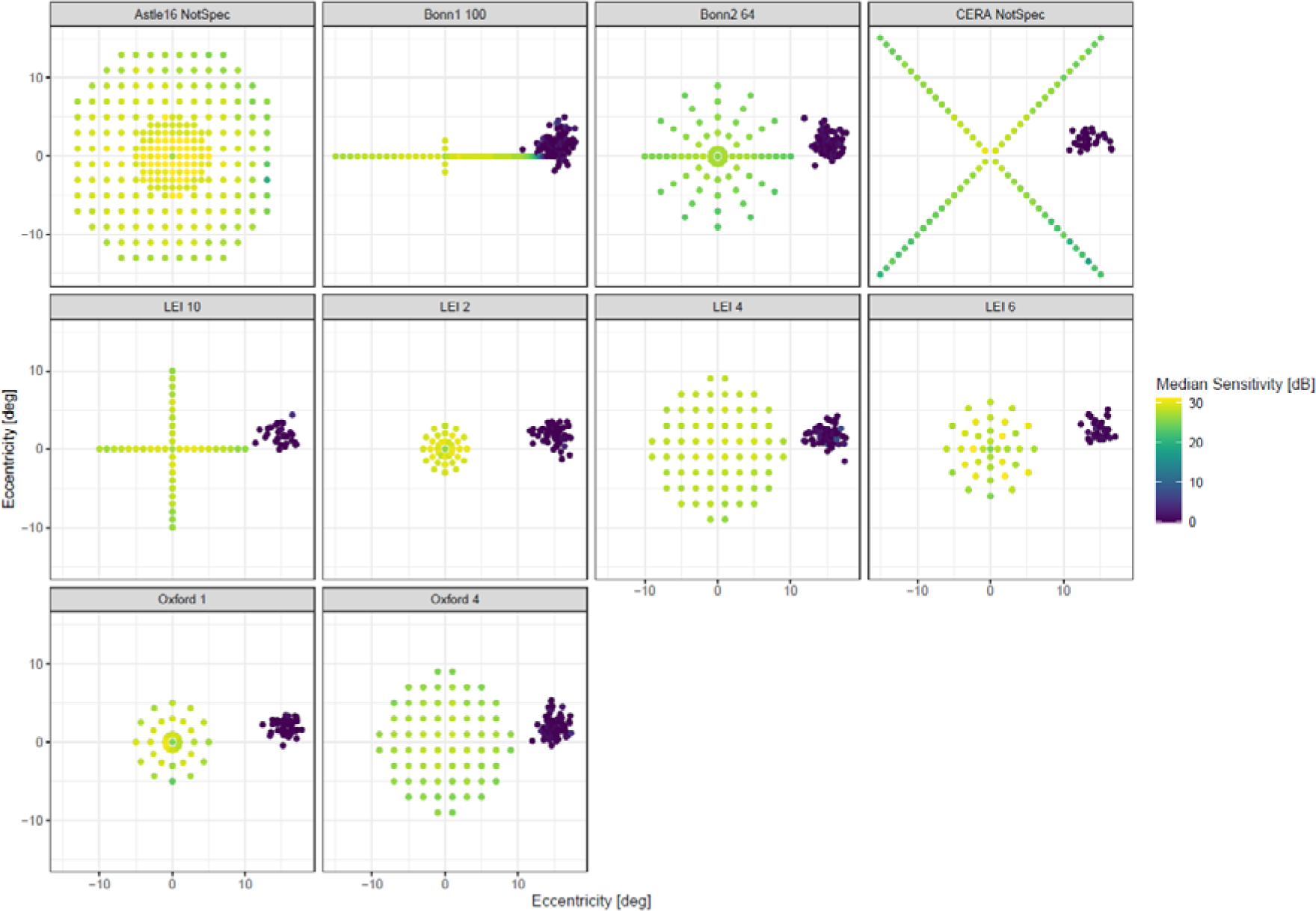
Included Perimetry Data. The dot plots show the perimetry data included in this study. The color denotes the sensitivity. The ‘0 dB’ dots on the temporal margin of the image denote the position of the optic nerve head.

**Table 1.** Included Data.

|  | Participants | Age (Median [Min, Max]) | Female | Male | Grid | Eyes | N of Exams (Median [Min, Max]) |
| --- | --- | --- | --- | --- | --- | --- | --- |
| Astle16 | 60 | 23.0 [19.0, 50.0] | 38 (63.3%) | 22 (36.7%) |  | 60 | 1 |
| Bonn1 | 54 | 41.2 [19.1, 82.0] | 37 (68.5%) | 17 (31.5%) |  | 103 | 1.00 [1.00, 2.00] |
| Bonn2 | 102 | 59.4 [18.0, 84.5] | 38 (37.3%) | 64 (62.7%) |  | 102 | 2.00 [1.00, 2.00] |
| CERA | 28 | 69.0 [53.0, 79.0] | 16 (57.1%) | 12 (42.9%) |  | 33 | 3.00 [1.00, 3.00] |
| LEI | 103 | 52.0 [16.0, 75.0] | 49 (47.6%) | 54 (52.4%) |  |  |  |
|  | 32 | 62.0 [21.0, 75.0] | 16 (50.0%) | 16 (50.0%) | LEI 10 | 32 | 1.00 [1.00, 1.00] |
|  | 58 | 53.5 [21.0, 75.0] | 26 (44.8%) | 32 (55.2%) | LEI 2 | 65 | 1.00 [1.00, 3.00] |
|  | 63 | 55.0 [16.0, 75.0] | 24 (38.1%) | 39 (61.9%) | LEI 4 | 74 | 1.00 [1.00, 2.00] |
|  | 20 | 45.0 [20.0, 74.0] | 14 (70.0%) | 6 (30.0%) | LEI 6 | 34 | 1.00 [1.00, 1.00] |
| Oxford | 85 | 29.0 [18.0, 85.0] | 45 (52.9%) | 40 (47.1%) |  |  |  |
|  | 47 | 29.0 [18.0, 76.0] | 27 (57.4%) | 20 (42.6%) | Oxford 1 | 48 | 7.00 [1.00, 13.0] |
|  | 40 | 37.0 [19.0, 85.0] | 19 (47.5%) | 21 (52.5%) | Oxford 4 | 80 | 1.50 [1.00, 3.00] |
| **Overall** | **432** | **45.0 [16.0, 85.0]** | **223 (51.6%)** | **209 (48.4%)** |  | **N=531** | **1052** |

**Note:** Some participants at LEI and Oxford were tested with multiple grids. Thus, the total number of participants is smaller than the number of participants per grid.

Supplementary Table S2. shows an overview of the eligibility criteria of the studies. Microperimetry testing was performed in all studies using the MAIA device. The background luminance was mesopic (1.27 cd/m^2^) and the device offered a dynamic range of 36 dB. All studies used the pre-set 4-2 staircase strategy (stimulus size: Goldmann III, stimulus duration 200 ms).

### Data Extraction and Availability

Sensitivity data was extracted from the *.tgz export of the MAIA device using a custom R script. All data is available at *[Zenodo repository, link will be provided upon acceptance]*. An *R* vignette to obtain personalized normative data by using limited site-specific data in combination with the now published data will be available at: *[Zenodo repository, link will be provided upon acceptance]*.

### Data Analysis

The analysis was structured in three parts: (1) fitting of parsimonious (interpretable) hill-of-vision models, (2) systematic comparison of regression algorithms for centile estimation regarding their goodness-of-fit and coverage, (3) analysis of the benefit of site-specific data regarding goodness-of-fit and coverage.

#### Parsimonious Hill-of-Vision Models

To obtain a parsimonious/interpretable hill-of-vision model, we fitted a linear mixed models (LMMs) using the *R* package *lmer*. In this null model, the hill-of-vision was described as a cone (linear effect of eccentricity), that could be shifted and up- and downwards (linear effect of age). In addition, we added a binary term indicating overlap with the fixation target based on previous literature (i.e., ‘foveal’ for eccentricities <1°).^21^ In the Wilkinson-Rogers syntax, the null model was:

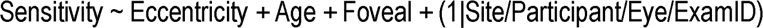

We also explored more complex models (Supplementary Results 1) selected by stepwise forward selection based on the Bayesian information criterion (BIC). Based on previous hill-of-vision modeling,^22^ we considered the following additional explanatory variables: eccentricity^2^ (quadratic term to describe steepening/flattening of the peripheral hill-of-vision), age^2^ (quadratic term describe steepening/flattening of the effect of age), *X coordinate + Y coordinate* (plane to tilt the hill-of-vision), superior-eccentricity + inferior-ecc. + temporal-ecc. + nasal-ecc. (pyramidal deformation of the hill-of-vision), eccentricity × age (interaction to describe variation in the ageing effect in dependence of eccentricity), sex, baseline exam (binary indicator to differentiate baseline exams from follow-up exams).

#### Comparison of Regression Models and Prediction Interval Calibration

The regression algorithms for centile estimation were compared using 6-fold cross-validation (site-wise splits). Thus, the resulting goodness-of-fit and coverage estimates are representative for the performance at a future (‘unknown’) clinical site. For models requiring hyperparameter optimization (e.g., LASSO regression), an additional layer of 10-fold cross-validation was nested within each outer fold.

#### Candidate models

To model the average differential light sensitivity and quantiles, we considered both complex and parsimonious (i.e., interpretable) models.

- **Linear regression**: For the linear regression models, we used the prediction intervals to obtain the quantiles. The sequence of models was based on the results from the step-wise forward selection in the section above (see also Table 2).
  `o` LM0: Sensitivity ∼ Ecc. + Age + Foveal
  `o` LM1: LM0 Features + Superior-Ecc. + Inferior-Ecc. + Temporal-Ecc. + Nasal-Ecc.
  `o` LM2: LM1 Features + Eccentricity×Age Interaction
  `o` LM3: LM2 Features + Eccentricity^2^
  `o` LM4: LM3 Features + Age^2^
- **Quantile regression**: For quantile regression, we fitted the formula from LM1 (to allow for variation of the centile surface across the visual field) using the R package *quantreg*.^23^
- **Penalized quantile regression with LASSO** For LASSO regression, we added all explanatory variables and their two-way interactions as candidate features and standardized all data (standardization performed within each analysis set and then applied to the respective assessment set). Within each analysis set, we performed 10-fold cross-validation to select the optimal L1 penalty for each of the quantile of interest (0.1, …, 0.9), employing the *R* package *rqPen*.^24^
- **Bayesian nonparametric quantile regression** The model included an isotropic smooth (thin plate regression spline) for the x and y coordinate, a spline for the age and a tensor product smooth for age and eccentricity. We used the R package *qgam* to obtain the quantiles of interest (0.1, …, 0.9).^25^
- **Bayesian mixed model** We fitted Bayesian mixed models with the above formula from LM1 (to allow for variation of the centile surface across the visual field) and with the clinical site and participant as random effects using the R package *brms*.^26^ The predict intervals function included the group-level uncertainty for new data based on the variation among the existing levels clinical sites and participants.

**Table 2.**
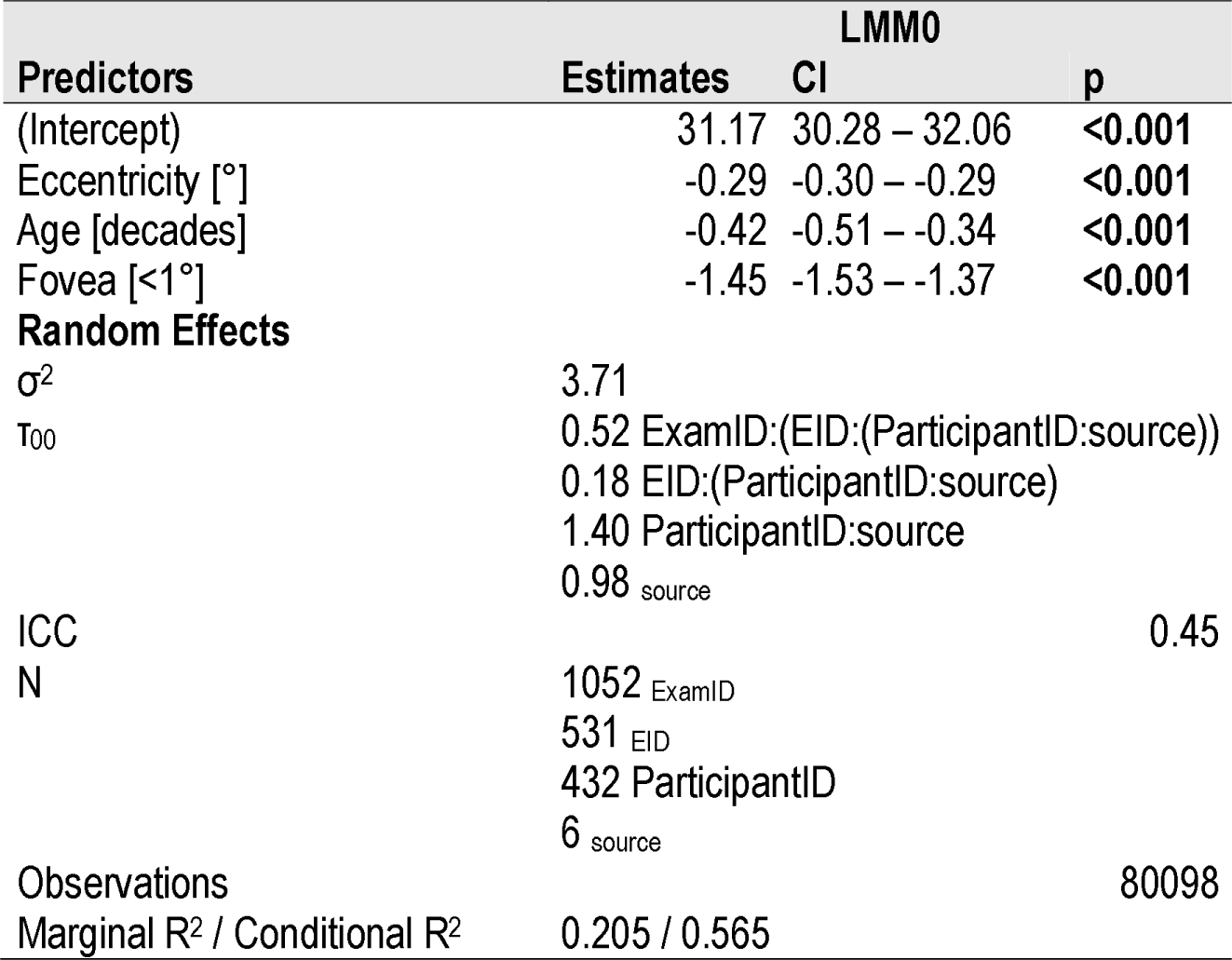
Linear Mixed-Model Analysis.

## Performance Metrics

To compare the models, we considered four factors. These concepts are visually explained in Figure 2.

**Figure 2.**
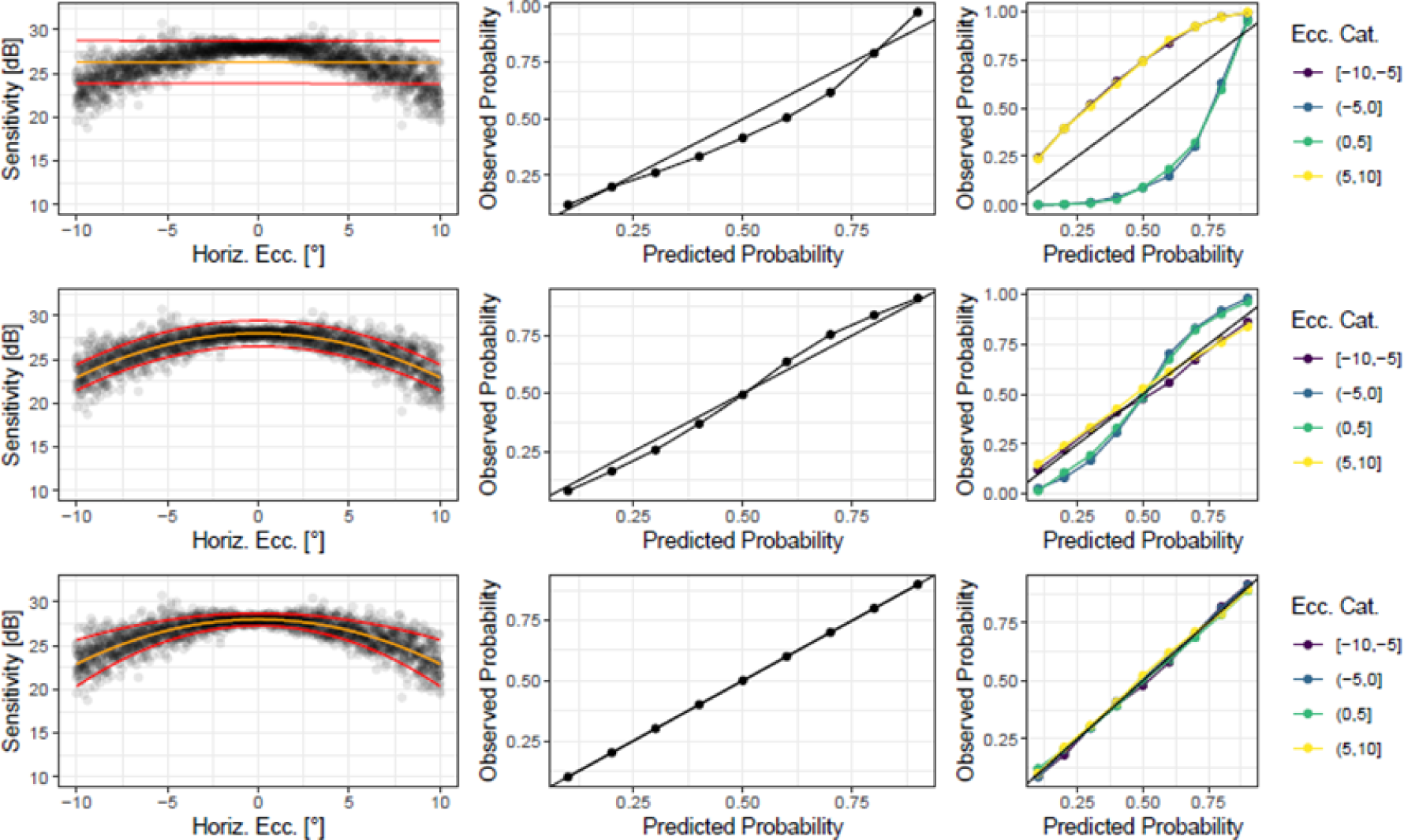
Concepts. The figure explains key consideration for the assessments of normative data, illustrated with simulated data. The left column shows the prediction (orange line) and the 80% prediction interval (red lines). The middle column shows the calibration plots across all data. The right column shows the calibration plots stratified by eccentricity. The miscalibration area (i.e., area between the calibration lines and the ideal diagonals) is reflective of the model calibration. The first model (row 1) is unsuitable to describe the data due to a poor goodness-of-fit. The second model (row 2) fits the data well and shows good overall calibration, but the prediction interval is constant resulting in poor conditional calibration (i.e., poor adaptivity). The third model (row 3) shows excellent goodness-of-fit, overall calibration, and conditional calibration.

- **Goodness-of-fit** The mean absolute error (MAE in dB) between measured sensitivity and predicted sensitivity with lower values indicating better performance.^27^
- **Coverage (marginal coverage)** The prediction intervals should cover the pre-specified fraction of measurements across locations. The coverage can be visualized by plotting the observed against the predicted proportion of measurements in the interval and quantified based on the mean absolute calibration error (MACE).^28,29^
- **Adaptivity (conditional coverage)** Prediction intervals should be smaller in easier-to-predict regions in the feature space (e.g., young individuals) and larger prediction in challenging-to-predict regions in the feature space (e.g., older individuals). As a metric of adaptivity, we assessed the MACE stratified by age and eccentricity bins.^28,29^
- **Parsimony** Regarding complexity of the computation and interpretability, parsimonious models are preferable.

As measure of goodness-of-fit, we estimated the MAE using linear mixed models (fitted with *lme4*) by estimating the absolute error (between predicted and measured sensitivity) for each test point with the regression model as the explanatory variable. The exam nested in eye, nested in participant, nested in site was considered as a random effects term. As measure of calibration, we computed the miscalibration area (i.e., absolute area between the calibration line and the ideal diagonal, cf. Figure 2).

## Effect of Site-Specific Data

Last, we used repeated (5 repeats) 5-fold cross validation with per-participants splits to assess the performance of the best-performing normative data model (Bayesian mixed model) with site-specific training data.

In addition, we implemented a simulation analysis to assess, how much site-specific data is needed to obtain a reliable model fit. For each clinical site, we fitted a normative data model with the data from all but one site, adding in a step-wise manner 2, 4, 8, 16 local participants. The performance of the resulting normative data model was then assessed on remaining participants of this site. We repeated this procedure 25 times. In this simulation, we used LM0 with the site added as an additional explanatory variable. The parsimonious LM0 was elected in this simulation analysis considering computational burden.

## RESULTS

### Included data

This study included perimetry data from a total of 1,052 tests from 531 eyes of 432 healthy participants, who had a median age of 45 years [min: 16, max: 85]. Table 1 lists the details about the age, sex, and perimetry grids separately for each clinical center. Figure 1 and Supplementary Table S1 provide a detailed description of the perimetry grids.

### Modeling Mean Visual Sensitivity

Qualitative inspection of the data (Figure 3) revealed that eccentricity and age were associated with visual sensitivity. Sensitivity was slightly lower in proximity to the fixation target (at eccentricities <1°), peaked at 2°, and then decreased with increasing eccentricity.

**Figure 3.**
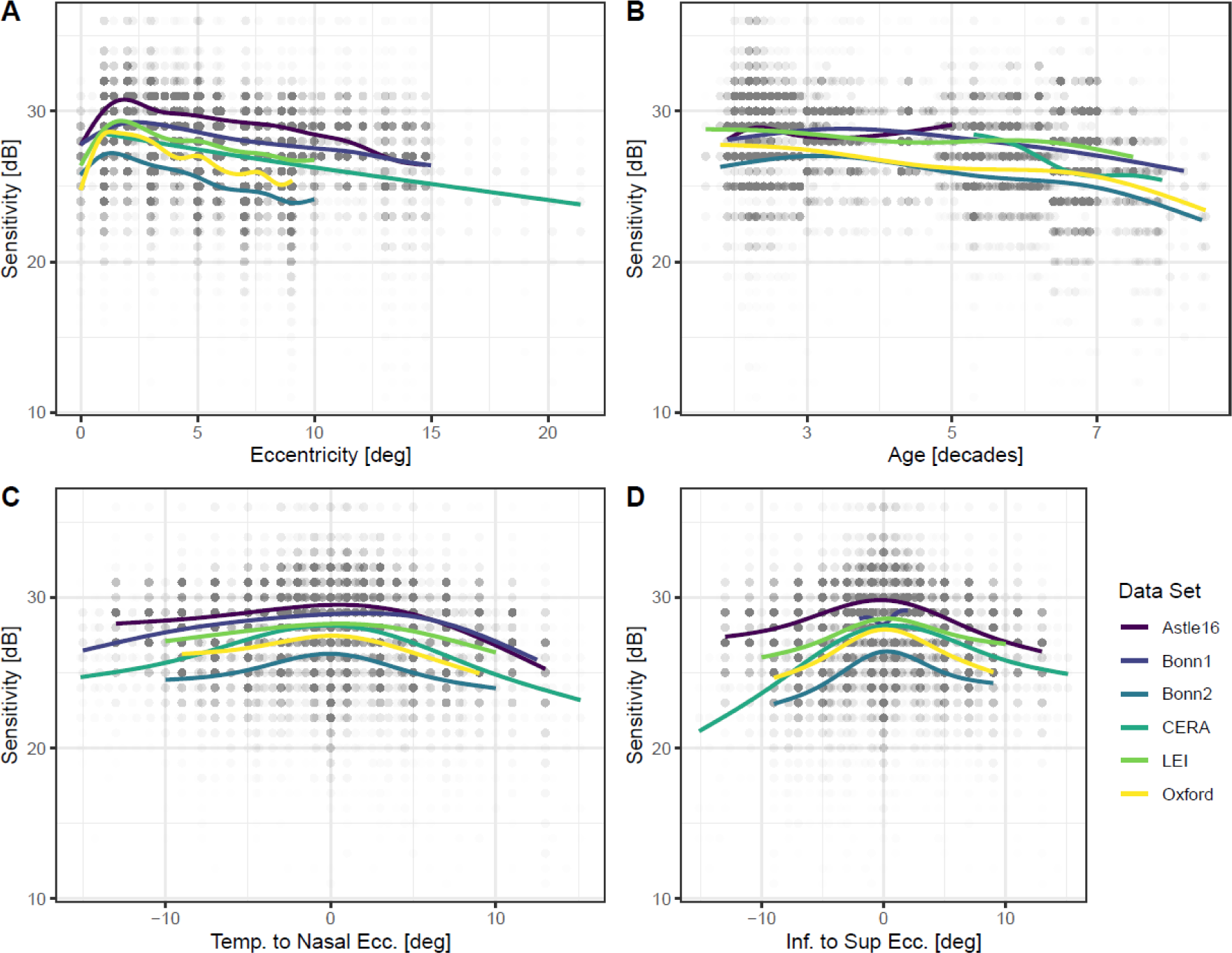
Visual Sensitivity. The panels show retinal sensitivity in dependence of eccentricity (A), age (B), position along the horizontal (C), and vertical (D) meridian. The colored lines are regression splines. Besides the dip in sensitivity in proximity to the fovea (fixation target), mesopic sensitivity decreases linearly with eccentricity as a first order approximation in the central visual field.

The null model (LM0, linear mixed model to predict sensitivity with eccentricity, age and foveal location as predictors) had a substantial explanatory power (*conditional R*^2^ of 56.5%) with a *marginal R*^2^ relating to the fixed effect alone of 20.5% (Table 2). The model’s intercept was 31.17 dB (95% CI) [30.28, 32.06], the effect of eccentricity was −0.29 dB/° [-0.30, −0.29], and the effect of age was −0.42 dB/decade [-0.51, −0.34]. The foveal test point in proximity to the fixation target (i.e., eccentricity <1°) was predicted to have a lower sensitivity (−1.45 dB [−1.53, −1.37]).

The random effects estimates (random intercept variance [τ00]) revealed considerable between-site/device variability (SD = √(0.98 dB^2^) = 0.99 dB) and between-participant variability (SD = √(1.40 dB^2^) = 1.18 dB). The between-eye variability was small (SD = √(0.18 dB^2^) = 0.42 dB), and there was within eyes evidence of exam-to-exam variation (SD = √(0.52 dB^2^) = 0.72 dB).

Using stepwise forward selection allowed to fit LMMs with a slightly better goodness-of-fit, but the improvements in marginal *R*^2^ were small (Supplementary Table S3).

### Comparison of Normative Data Models

Overall, most regression models performed similar data from ‘new’ clinical sites (i.e., cross-validation with the clinical site as the grouping variable). The MAE between predicted and observed sensitivity ranged from (mixed model estimate [95% CI]) 2.13 dB [1.86, 2.4] for the Bayesian mixed model to 2.24 dB [1.97, 2.51] for the linear model 2 (Figure 4A, Table 3). Post-hoc pair-wise comparison of the MAE revealed that the Bayesian mixed model performed indeed statistically significantly better compared to the other models, although the actual magnitude of difference was small (Supplementary Table S4).

**Figure 4.**
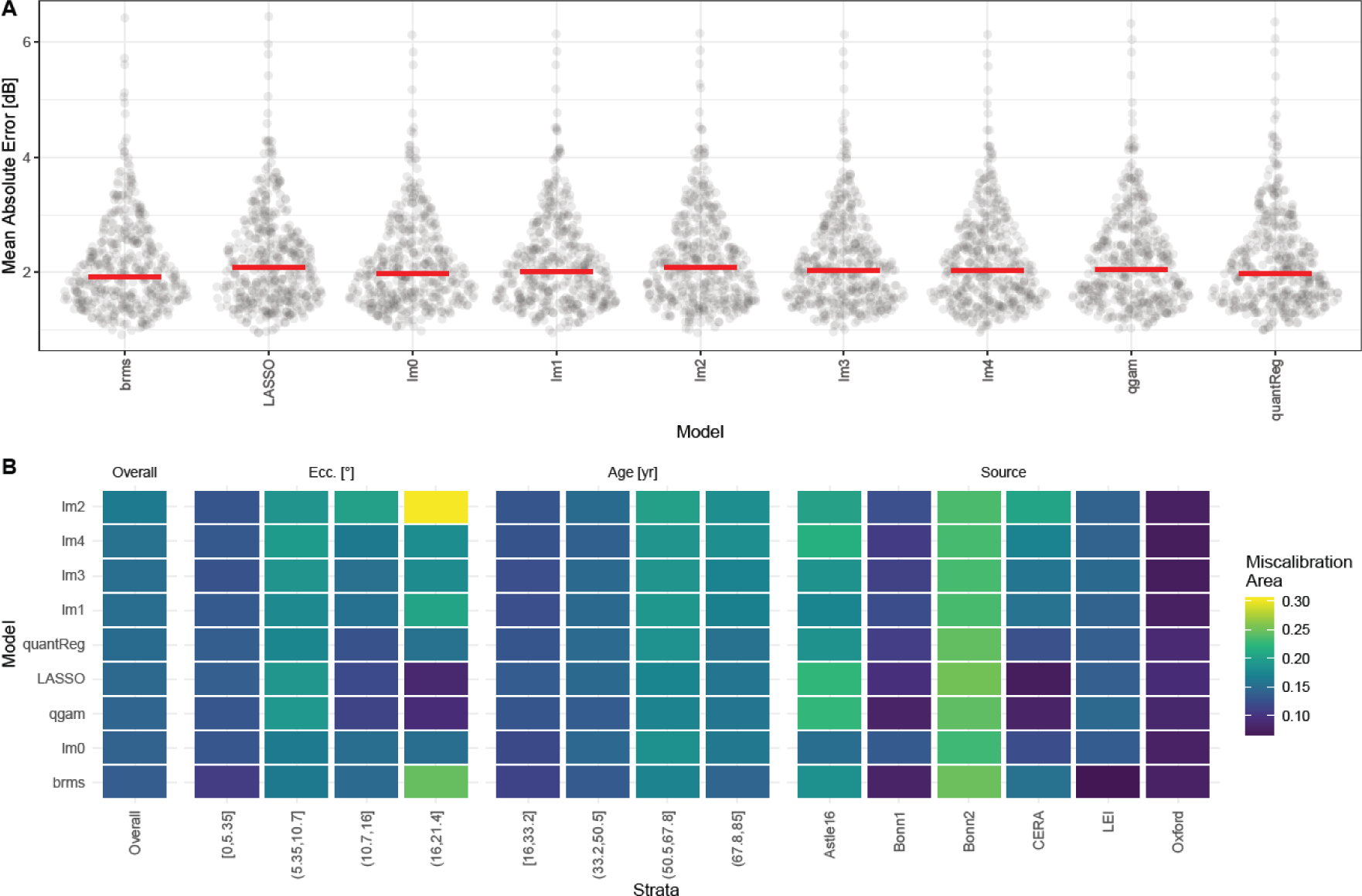
Cross-Validated Model Perfromance with Site-Wise Splits. Panel A shows the mean absolute error between predicted and observed sensitvity (each dot represents one participant, the red bar shows the median). Panel B shows the miscalibration area overall, and stratified by eccentrictiy (ecc.), age, and clinical site. The overall miscalibration area is remarkably similar across models. The Bayesian mixed model had the smallest miscalibration area. The Bayesian non-parametric additive model and LASSO model generalized the best to data with eccentic test points (i.e., eccentrictiy >10.7°). *Abbreviations: Bayesian mixed model (brms), Linear model (lm), Bayesian non-parametric additive model (qgam), Quantile Regression (quantReg)*

**Table 3.**
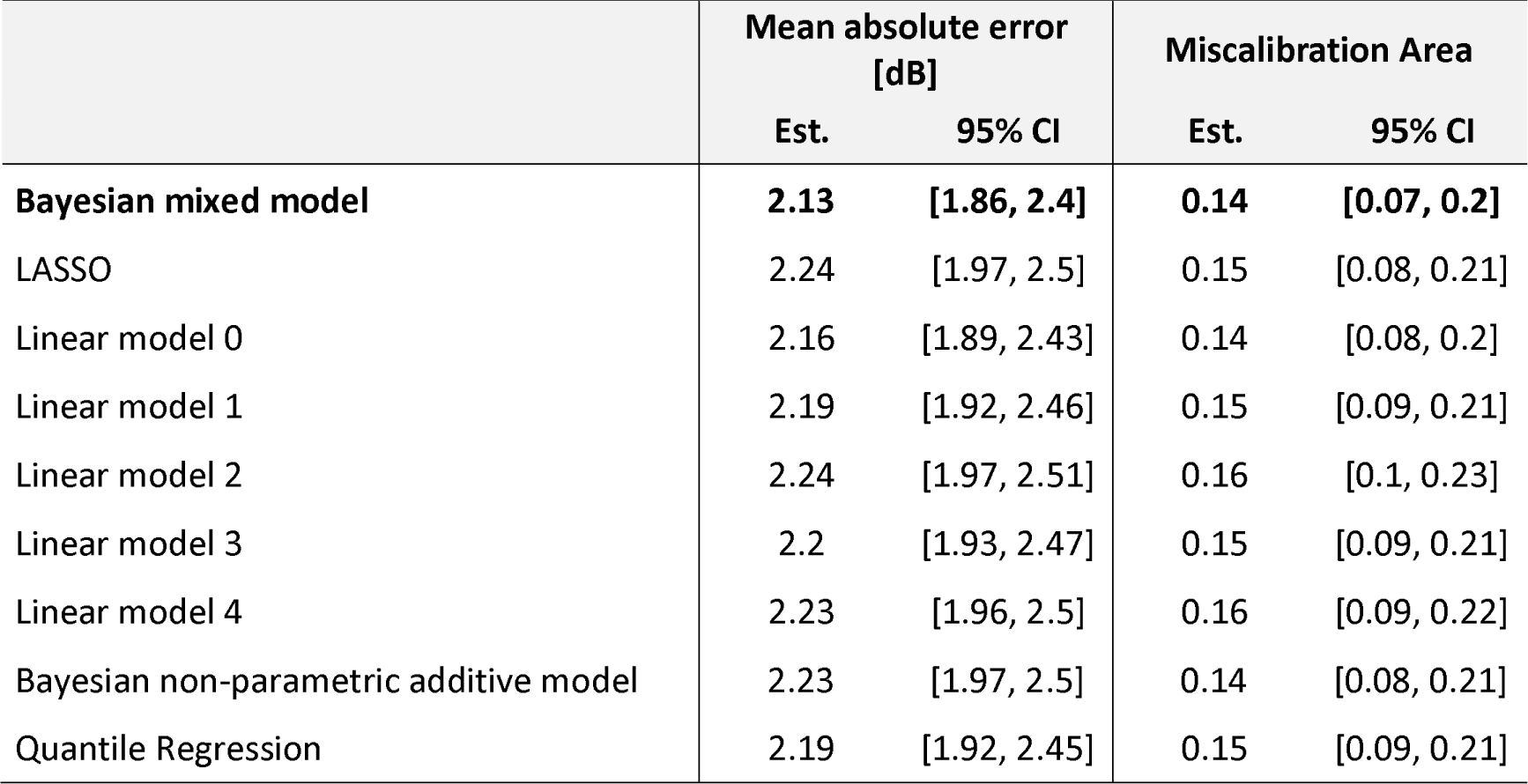
Model Performance with Cross-Validation by Clinical Site.

In terms of overall calibration (marginal coverage), the Bayesian mixed model also showed the best performance, with a miscalibration area of 0.14 [95% CI = 0.07, 0.2] (Table 3). But the calibration curves stratified by eccentricity (conditional coverage) indicated that for the Bayesian non-parametric additive model and LASSO regression models performed better in test points with >10.7° eccentricity (Supplementary Figure S2).

### Normative Data Models with Site-Specific Data

With cross-validation using participant-wise splits (and adding the clinical site as a predictor variable), the LASSO model performed best in terms of MAE (1.79 dB [1.65, 1.93]) and the Bayesian non-parametric additive model best in term of miscalibration area (0.03 [0.01, 0.05]) (Supplementary Table S5, Supplementary Figure S3). Post-hoc comparison of the models revealed that those to models performed comparably with regard to those metrics (i.e., no statistically significant differences, Supplementary Table S6). Again, all regression models were comparable for central test point, but the conditional coverage for test points with an eccentricity >10.7° was markedly better for the LASSO and Bayesian non-parametric additive model (Supplementary Figure S4). Figure 5 shows the visual field surfaces predicted by the Bayesian non-parametric additive model as a function of age.

**Figure 5.**
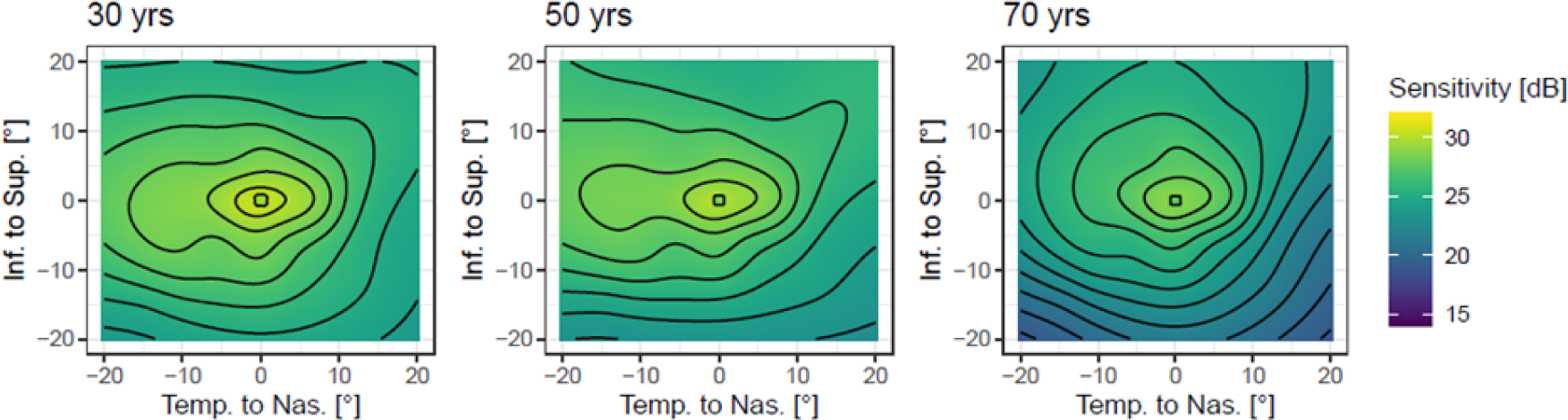
Effect of Ageing Visualized. The panels show the expected 50^th^ percentile surfaces for particpants at age 30, 50, and 70 years. The black lines denote 1 dB contour lines. Notably, the effect of aeging between 30 and 50 years is mild. Most ageing is observable later with a notable steepening of the decline in retinal senstivity along the inferior meridian in the mid-periphery.

### Impact of Adding Site-Specific Data

Since adding site-specific data drastically improves model calibration, we assessed how much site-specific data is needed for this gain. Thus, we evaluated the impact of adding limited site-specific data (e.g., 2, 4, 8 or 16 randomly selected participants) to the model fitting process. Adding site-specific data improved the model fit and calibration markedly (Supplementary Figure S5 and S6). Data from just eight site-specific participants was sufficient for achieving quasi-optimal fit and calibration (Supplementary Table S7).

### Effect of Follow-Up and Sex

Microperimetry tests are usually performed without prior information available at each specific location to seed the thresholding procedure during the first test (a “baseline” test), except for initial estimates obtained at four primary seeding locations.^7^ However, subsequent tests can use estimates from the previous test to seed its testing (which is usually referred to as a “follow-up” test). There was no significant difference in the final goodness-of-fit or calibration of the Bayesian non-parametric additive model based on whether a test was a follow-up or baseline test, or based on the sex of the participant, and these variables were thus not included in the above model fitting procedures (Supplementary Table S8).

## DISCUSSION

This study presents an extensive dataset for mesopic microperimetry collected from multiple centers. It compares quantile regression techniques based on their goodness-of-fit, marginal coverage, and conditional coverage, while also examining the usefulness of site-specific normative data. These data are crucial for analyzing microperimetry data, both for everyday clinical use and for research used, particularly in the setting of patient-tailored (individualized) perimetry and multicenter clinical trials.

Importantly, our models recapitulate free-viewing perimetry data on the mesopic hill of vision. Aulhorn and coworkers demonstrated that the shape of the mesopic hill-of-vision mirrors mostly the photopic hill-of-vision, but with a slightly flatter slope of the sensitivity decrease with increasing eccentricity.^30^ This shape was later recapitulated for photopic and mesopic perimetry studies using the Octopus perimeter and the Humphrey perimeter.^22,31,32^ Our parsimonious linear mixed model analysis for data from the MAIA perimetry was compatible with these prior data, with the exception of the relatively low foveal sensitivity. This peculiarity for the MAIA device was reported previously and is linked its fixation target close to the fovea.^21^ Our models are also in line with the previous quadratic trend surface-based model for the MAIA from Denniss and Astle with a relatively flat hill-of-vision in the temporal macula.

Regarding the underlying biology, spectral perimetry studies imply that Goldmann III stimuli in mesopic conditions are mediated by a combination of S-cones and M+L-cones centrally (<13°), while for more peripheral sensitivity is determined by rods and M+L-cones.^31^ Thus, our central data are compatible with the human photoreceptor topography considering the ellipsoid-shaped topography of the cone density in the central 10°.^33^ With seven decades of aging, light sensitivity will decrease by 3 dB based on our data. Thus, our data are further support by previous histopathologic studies which showed a 30.7% combined cone and rod density loss in the central 4 mm region with seven decades of aging (assuming that the square of photoreceptor density is proportional to light sensitivity in linear units).^34–36^

Apart from the central tendency of normal visual sensitivity, detailed knowledge about its expected healthy inter-individual and measurement variability is critical for detecting early dysfunction. First, we used linear mixed model to identify sources of variability. The largest source of variability was the retest variability, consisting of the point-wise residual variance and global fluctuations. The derived 95% repeatability coefficients for the point-wise sensitivity and the mean sensitivity (approx. ±6 dB and ±2 dB, respectively) are in line with previous retest reliability studies in healthy subjects and patients with retinal diseases.^37–39^ Moreover, these global fluctuations (i.e., vigilance, positioning in front of the device) are comparable to free-viewing perimetry estimates in the absence of severe visual field defects.^40,41^ In addition, there was considerable between-participant variability but only very little between eye variability. There was also site-associated variability (discussed in the next paragraph).

The regression algorithms for centile estimation performed all very similarly in terms of goodness-of-fit and marginal coverage (overall calibration). However, the Bayesian mixed model outperformed the other models in terms of conditional coverage (calibration in age- or eccentricity-strata) with site-wise cross-validation. The higher prediction error of the linear models is likely an indicator of overfitting, whereas the Bayesian mixed model is regularized through the priors and the partial-pooling.

Nevertheless, all regression models used in our study were not very effective in generalizing across different sites. Our study design does not provide information on whether the differences observed between sites are due to device calibration, test conditions, stimulus pattern used, or biological differences. However, a random intercept model that considers site-specific data achieved perfect calibration across sites (Supplementary Figure S6). Thus, device calibration-related differences are a plausible explanation. Based on the ISO standard for perimetry devices, the calibration tolerance for the increment luminance over the background (ΔL/LB) is +25 % to −20%.^42^ This corresponds to dB scale shifts of ±1 dB. All except one site random effects estimates for the site fell into this range.

Clinically, relying solely on the manufacturer’s normative data could result in diagnostic errors, particularly in older individuals and in test points located near the peripheral macula.^17,18^ The manufacturer defines sensitivity measurements below 25 dB as abnormal, based on data from 270 subjects with a mean age of 43 years ± 15.^43^ The Gaussian distribution shown on the pre-set printout indicates that measurements below 25 dB are outside of 3 standard deviations (i.e., 0.135% quantile), which is also reflected by the device’s pre-set color palette for displaying test results. However, our analysis and previous studies have revealed that aging and eccentricity have a significant impact on mesopic retinal sensitivity.^17,18^ Based on our findings, we predict for example that around half of the sensitivity measurements at 10° eccentricity will fall below 25 dB in healthy individuals at the age of 75.

For research, sensitivity loss should ideally be computed with site/device-specific normative data. Using the here published data and code vignette in *R* Markdown (*Github Link TBD*), site/device-specific normative data can be obtained efficiently with 8 to 16 normative subjects.

### Limitations

A key limitation of this study is that the data collected was retrospective in nature and performed using different stimulus patterns. In addition, the testing protocol (e.g., pupil dilation, or prior light exposure^44^) could differ among sites, as well as the definition of healthy individuals. Pupillary dilation has been empirically shown to have no effect (given the small required pupil diameter of 2.5 mm).^45^ Given that surgery-requiring cataract has only a 1-2 dB effect on visual sensitivity,^46^ the variability due to cataracts in some healthy individuals is likely small. Based on our data, no sex differences were evident. But large epidemiologic data implies small differences, that would necessitate a markedly larger sample size to detect.^47^

### Summary

In conclusion, our study provides a comprehensive analysis of normal retinal sensitivity measured with the MAIA device across different age ranges. Determinants of mesopic light sensitivity were the eccentricity and age. Considerable between-site variability was evident, which could be due to device calibration, participant population, or protocol differences.

## Data Availability

All data produced will be published at Zenodo.org upon acceptance of the manuscript.

## Commercial Relationships Disclosures

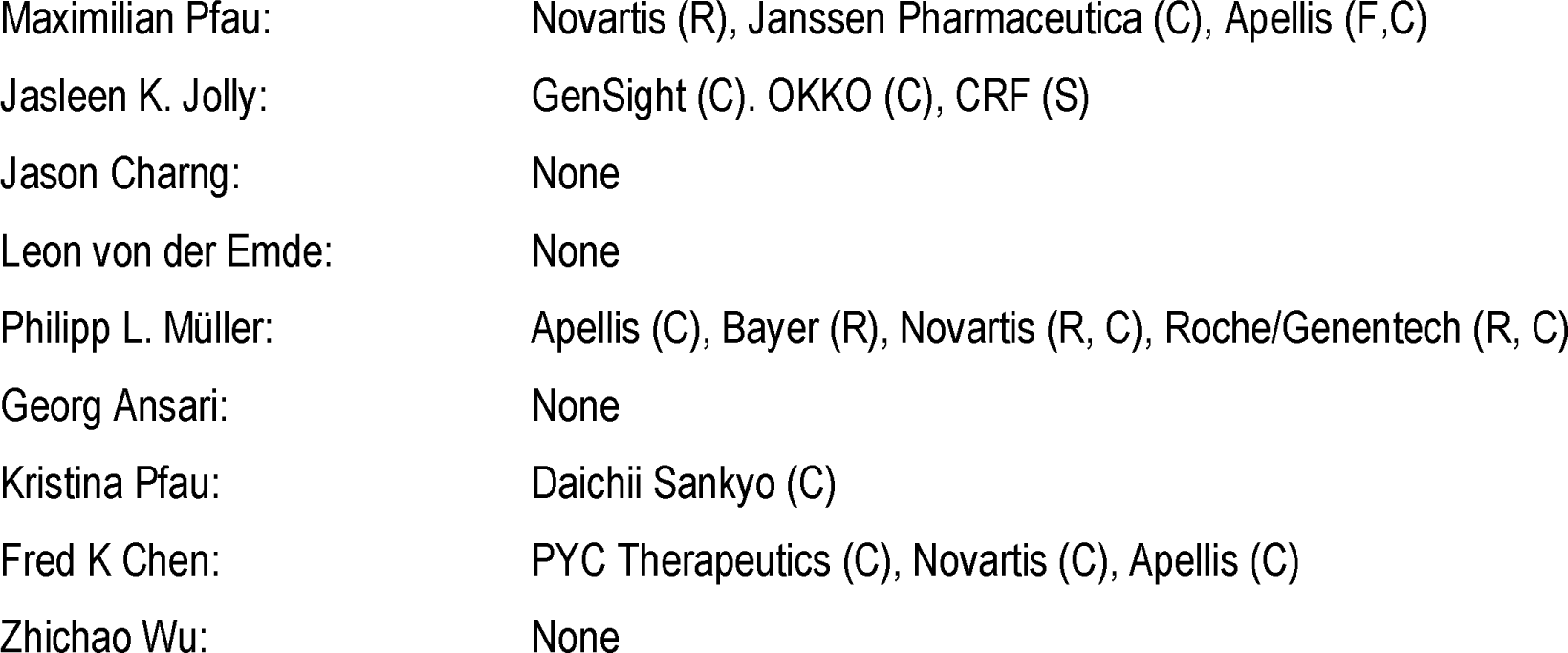

**Commercial Relationships Disclosures:**
|  |  |
| --- | --- |
| Maximilian Pfau: | Novartis (R), Janssen Pharmaceutica (C), Apellis (F,C) |
| Jasleen K. Jolly: | GenSight (C), OKKO (C), CRF (S) |
| Jason Charng: | None |
| Leon von der Emde: | None |
| Philipp L. Müller: | Apellis (C), Bayer (R), Novartis (R, C), Roche/Genentech (R, C) |
| Georg Ansari: | None |
| Kristina Pfau: | Daichii Sankyo (C) |
| Fred K Chen: | PYC Therapeutics (C), Novartis (C), Apellis (C) |
| Zhichao Wu: | None |

